# Measuring post-discharge socioeconomic and quality of life outcomes in trauma patients: A scoping review

**DOI:** 10.1101/2021.05.13.21257073

**Authors:** Siddarth David, Nobhojit Roy, Harris Solomon, Cecilia Stålsby Lundborg, Martin Gerdin Wärnberg

## Abstract

**Purpose:** Managing trauma is a global public health challenge. Measuring post-discharge socioeconomic and quality-of-life outcomes can help better understand and reduce the consequences of trauma.

**Methods:** We performed a scoping review to map the existing research on post-discharge outcomes for trauma patients, irrespective of the country or setting in which the study was performed. The scoping review was conducted by searching six databases: MEDLINE, EMBASE, the Cochrane Library, Global Index Medicus, BASE, and Web of Science to identify all articles that report post-discharge socioeconomic or quality of life outcomes in trauma patients from 2009 to 2018.

**Results:** 758 articles were included in this study, extracting 958 outcomes. Most studies (82%) were from high-income countries (HICs). More studies from low- and middle-income countries (LMICs) were cross-sectional (71%) compared with HIC settings (46%). There was a wide variety of different definitions, interpretations, and measurements used by various articles for similar outcomes. Quality of life, return to work, social support, cost, and participation were the main outcomes studied in post-discharge trauma patients.

**Conclusions:** The wide range of outcomes and outcome measures reported across different types of injuries and settings. This variability can be a barrier when comparing across different types of injuries and settings. Post-discharge trauma studies should move towards building evidence based on standardized measurement of outcomes.

## 1. Introduction

Every year around 4.5 million people die from trauma, which is more than the fatalities recorded from HIV/AIDS, tuberculosis, malaria, and maternal mortality combined [1, 2]. Globally, trauma accounts for one-tenth of disability-adjusted life-years (DALYs), a burden largely in low and middle-income countries (LMICs) [3, 4]. Improved trauma management has reduced the number of deaths and attention has shifted to non-fatal traumas [5]. Consequently, post-discharge outcomes among survivors is an emerging field in public health research [6–8].

Trauma can have social consequences, and can affect relationships, participation in the community, independent living, and social functioning [9, 10]. It could result in direct financial costs including out-of-pocket expenses for treatment as well as indirect financial costs due to disability or loss of livelihoods, pushing families into poverty [11, 12]. Some studies show that up to 40% of trauma survivors are unable to work even one year after discharge [13–15]. These socioeconomic outcomes together can have a substantial effect on the health-related quality of life (QoL) [16]. However, there is limited research on these long-term outcomes experienced by trauma survivors [17].

### 1.1 Rationale and Objectives

Given the limited understanding of the different socioeconomic and QoL outcomes among trauma patients across different populations and settings, collating the existing evidence base can highlight key findings and identify gaps [18, 19]. The aim of this paper is to provide an overview of the existing knowledge on post-discharge socio-economic and QoL outcomes in trauma patients. It will explore the different outcome measures and the tools used to assess these outcomes in post-discharge trauma patients.

## 2. Methods

### 2.1 Protocol and Registration

The protocol for this paper was developed using Arksey and O’Malley’s methodology for scoping reviews [20, 21]. It maps the extent and range of the field of study using five steps: (1) identifying the research question, (2) identifying relevant studies, (3) study selection, (4) charting the data, and (5) collating, summarizing, and reporting the results. The protocol was registered at Prospero before the review was undertaken (CRD42018103940) [22]. The Preferred Reporting Items for Systemic Reviews and Meta-Analyses extension for Scoping Reviews (PRISMA-ScR) Checklist was followed for reporting the results of the study (Supplementary Materials Table S1) [23].

### 2.2 Eligibility Criteria

Studies on post-discharge trauma patients reporting measures of socio-economic and QOL outcomes were included in this review. The outcomes were based on the International Classification of Functioning, Disability and Health (ICF) checklist [24] and the WHO Manual for estimating the economic costs of injuries due to interpersonal and self-directed violence (WHO Manual) [11]. They included: social functioning, social adjustment, community participation, social relationships, social behavior, independent living, loss of livelihood, return to work, out-of-pocket expenses, indirect financial costs, health expenditures, cost analysis, and socioeconomic factors. Publications in English using any design for measuring or reporting on these outcomes were included. We chose literature from September 2009 to August 2018 as this covered research that was relatively recent.

### 2.3 Information Sources and Search Strategy

To be comprehensive we used both published and grey literature sources. The published studies were searched using the electronic databases MEDLINE, EMBASE, and the Cochrane Library. For the grey literature, Global Index Medicus, BASE, and Web of Science were searched to identify reports and other articles. The search terms were developed based on ICF and the WHO Manual, and on inputs from the authors. The final terms were searched as keywords in the titles, abstracts and subject headings (like MeSH) as appropriate. The search strategy is available in Supplementary Material.

### 2.4 Selection of Sources of Evidence

While the criteria for selection were determined at the outset, certain criteria were defined post hoc as we became familiar with the literature. These were applied to all the articles. We excluded citations that involved injuries such as whiplash, needle-prick, aneurysms, neuropathies, and sexual assault. We excluded physiological, functional, and psychological outcomes. The selected studies were imported into the online systematic review software Rayyan [25].

The study selection process involved two stages of screening: first, a review of the title and abstract; and second, a full-text review. In the first stage of screening, SD and MGW independently screened the title and abstract of all retrieved results for inclusion. Any articles that were deemed relevant by at least one reviewer were included in the full-text review. SD and MGW independently screened full-text articles, and any discordance was reviewed once again by each reviewer until consensus was reached.

### 2.5 Data Charting Process, Data Items and Synthesis of Results

For the included articles, a data collection instrument was developed to capture the article’s relevance and to extract the outcomes studied. It included: publication year, study design, country, income setting (high, middle, low income country based on the World Bank classification [26]), type of trauma such as mechanism of injury (fall, assault, road traffic crash, etc.), the socio-economic and QoL outcomes, unit of measurement, and the follow-up period. The WHO Disability Assessment Schedule (WHODAS 2.0), which is based on the ICF framework, was used to classify the outcomes [27]. Of its six domains, mobility, self-care, getting along, life activities, and participation were used for classifying the outcomes extracted. In addition, two more domains were used: costs and quality of life.

### 2.6 Critical Appraisal of Quality of Results

No appraisal of methodological quality or risk of bias of the included articles was done. This is consistent with guidance on conducting scoping reviews [20, 23].

## 3. Results

### 3.1 Selection and Characteristics of Sources of Evidence

The initial search from all the sources generated 11,160 references, out of which 2,086 were selected based on their titles and abstract. After having read the articles in full, 758 articles were selected for inclusion in the review. The main reasons for excluding full-text articles were wrong outcome of interest (n=893) or wrong population of interest (n=316). The process flowchart is given in Figure 1.

**Figure 1.**
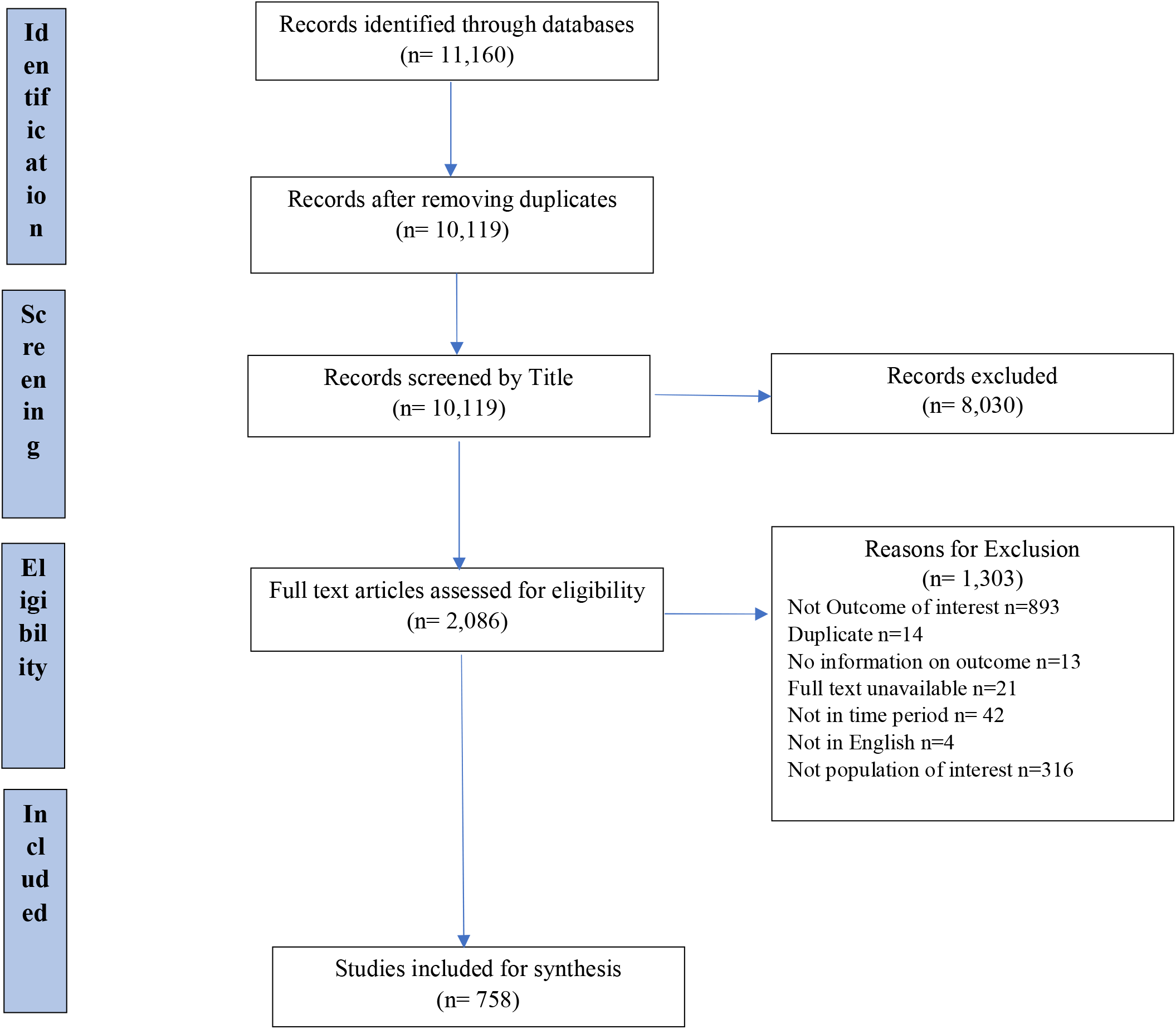
Flow of Studies According to PRISMA Flow Chart

Around one-tenth of the articles were unpublished reports (grey literature) such as conference abstracts, policy briefs, doctoral thesis, poster presentations, and consensus meetings (n=68). Of the remaining articles (n=690), 57 were reviews, including systematic reviews (n=37), 40 were qualitative, 18 were study protocols and tool validation studies, and nine were case series, editorials, or letters to editors. There were four articles that were analyses of secondary data or databases, and there was one modelling paper.

Out of the quantitative studies (n=561), a majority were cross-sectional or observational studies (n=302) followed by cohort-based studies (n=244). A majority of the cohort studies were prospective (n=207); the rest included longitudinal (n=15), retrospective (n=15), randomized control trials (n=5), and quasi-experimental studies (n=2). There were three articles that were retrospective analyses of prospectively collected data. Seven articles were mixed-method studies. Five of the articles were abstracts for which the full-text could not be found.

A majority (n=625, 82.5%) of the articles were from high-income countries, with the US (n=162), Australia (n=97), and the Netherlands (n=52) the top contributors. Of the 105 articles from middle-income countries (both upper and lower), China (n=14), India (n=14), and Iran (n=14) were the top contributors. Among the six articles from low-income countries, there were two from Nepal and one each from Benin, Haiti, Malawi, and Uganda. Of the articles, 22 were studies involving countries of different income levels.

There was a difference seen when comparing the study designs of articles by the income level of countries. Out of the 111 studies from LMICs, more than two-thirds (70.2%) had a cross-sectional design (n=78). The rest were cohort-based studies. On the other hand, of the 625 articles from HICs, only one-third (34.4%) were cross-sectional studies (n=215).

Of the articles that reported a follow-up period (n=574), the average follow-up was 56 months (around 4.5 years; the range was 0.25 months to 720 months). However, the follow-up period for most studies was up to one year (48.7%, n=280). Just over 12% of the studies had a follow-up period of more than five years (n=133).

In this review we divided the studies into two broad groups: studies on patients with a specific type of body injury (isolated injuries or with associated injuries) and studies on patients with a specific mechanism of injury. Most of the studies were based on patients with a specific type of body injury (n=604). A majority of the studies focusing on a specific body part were on traumatic brain injury (TBI) (n=262), followed by spinal-cord injury (n=171), limb injury (n=33), hip injury (n=18), and others (n=14). There were also 106 articles focusing on multiple body regions injured or polytrauma.

The remaining 154 articles were focused on patients with specific mechanisms of injury. The most common were burns (n=43) and road traffic injuries (n=43), followed by occupational injuries (n=31), sports injuries (n=14), and assaults (n=14). The rest included self-harm, falls, and terrorism (n=9).

### 3.2 Synthesis of Results

A total of 78 different outcomes were reported 982 times in the 758 studies included in the review. This is because many studies focused on more than one outcome. Classification of the extracted outcomes into domains is shown in Table 3.

**Table 1.**
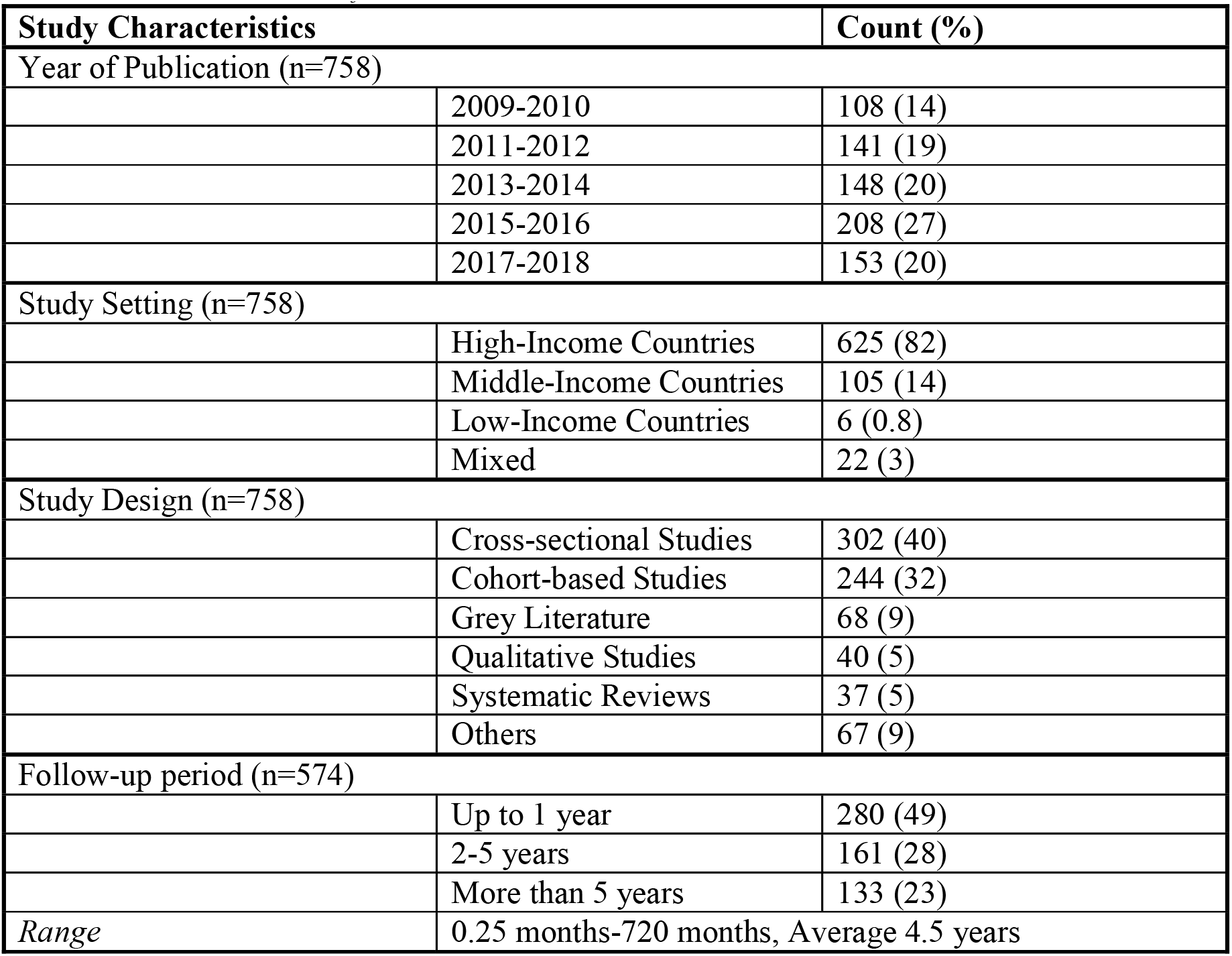
Characteristics of the Included Articles

**Table 2.** Types and Mechanisms of Injury in the Included Articles

| Type of Injury |  | Count (%) |
| --- | --- | --- |
| Body part injured (n=604) |  |  |
|  | Traumatic Brain Injury | 262 (43) |
|  | Spinal Cord Injury | 171 (28) |
|  | Polytrauma | 106 (18) |
|  | Limb Injury | 33 (6) |
|  | Hip injury | 18 (3) |
|  | Others | 14 (2) |
| Mechanism of Injury (n=154) |  |  |
|  | Road Traffic Injury | 43 (28) |
|  | Burns | 43 (28) |
|  | Occupational Injuries | 31 (20) |
|  | Sports injuries | 14 (9) |
|  | Assaults | 14 (9) |
|  | Others | 9 (6) |

**Table 3.**
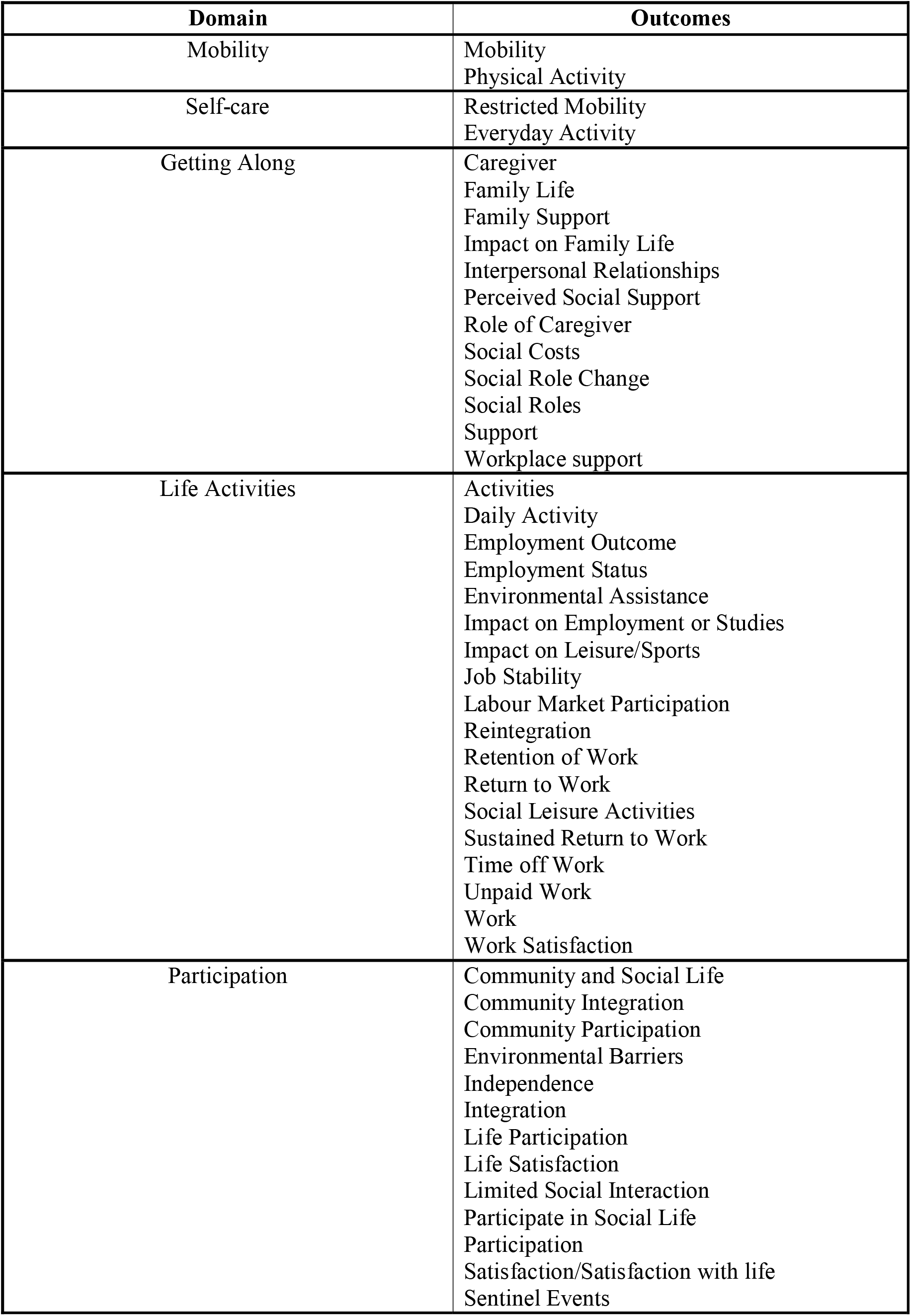
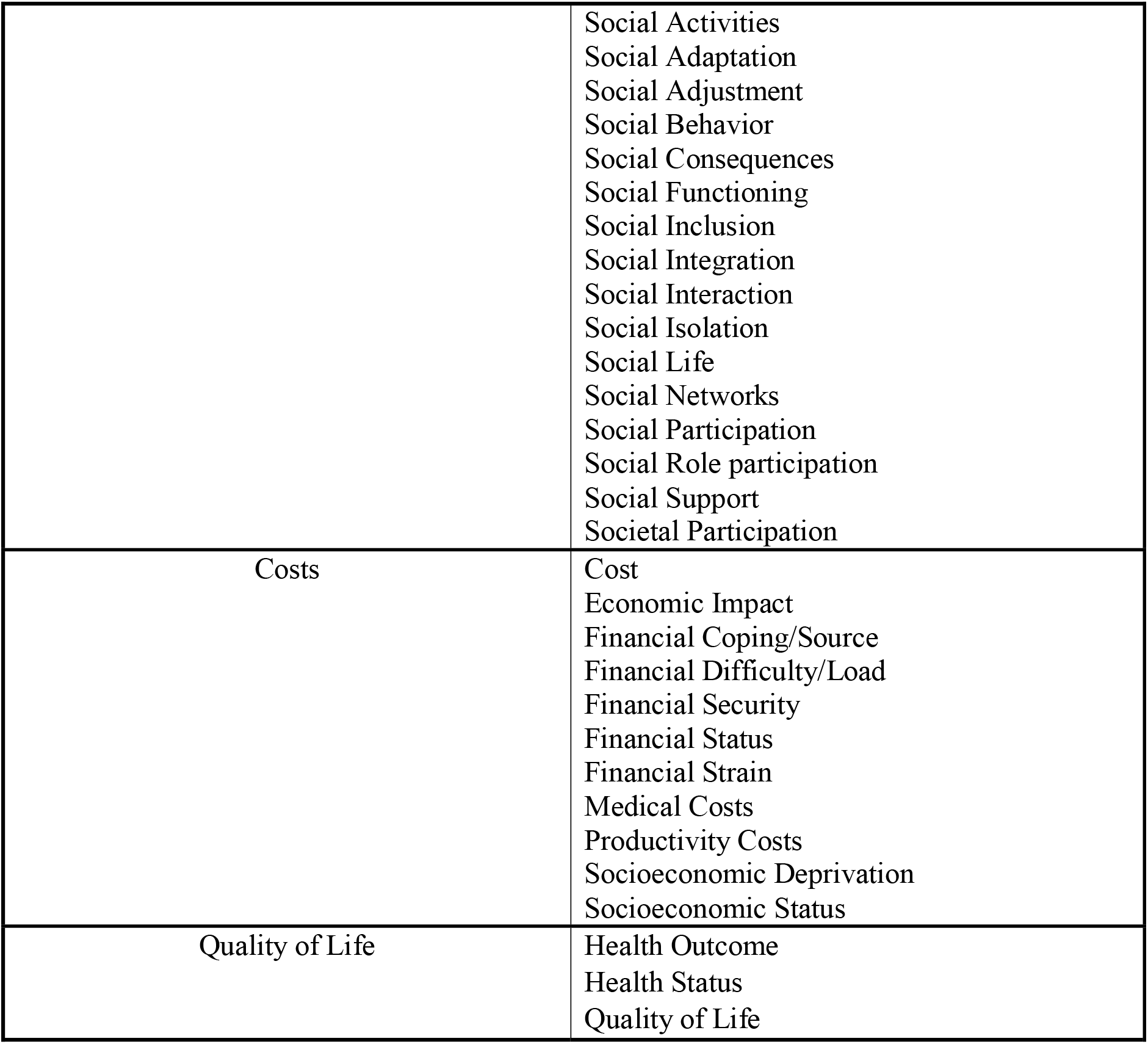
Classification of Reported Outcomes Into Domains

| **Domain** | **Outcomes** |
| --- | --- |
| Mobility | Mobility<br>Physical Activity |
| Self-care | Restricted Mobility<br>Everyday Activity |
| Getting Along | Caregiver<br>Family Life<br>Family Support<br>Impact on Family Life<br>Interpersonal Relationships<br>Perceived Social Support<br>Role of Caregiver<br>Social Costs<br>Social Role Change<br>Social Roles<br>Support<br>Workplace support |
| Life Activities | Activities<br>Daily Activity<br>Employment Outcome<br>Employment Status<br>Environmental Assistance<br>Impact on Employment or Studies<br>Impact on Leisure/Sports<br>Job Stability<br>Labour Market Participation<br>Reintegration<br>Retention of Work<br>Return to Work<br>Social Leisure Activities<br>Sustained Return to Work<br>Time off Work<br>Unpaid Work<br>Work<br>Work Satisfaction |
| Participation | Community and Social Life<br>Community Integration<br>Community Participation<br>Environmental Barriers<br>Independence<br>Integration<br>Life Participation<br>Life Satisfaction<br>Limited Social Interaction<br>Participate in Social Life<br>Participation<br>Satisfaction/Satisfaction with life<br>Sentinel Events |
|  | Social Activities<br>Social Adaptation<br>Social Adjustment<br>Social Behavior<br>Social Consequences<br>Social Functioning<br>Social Inclusion<br>Social Integration<br>Social Interaction<br>Social Isolation<br>Social Life<br>Social Networks<br>Social Participation<br>Social Role participation<br>Social Support<br>Societal Participation |
| Costs | Cost<br>Economic Impact<br>Financial Coping/Source<br>Financial Difficulty/Load<br>Financial Security<br>Financial Status<br>Financial Strain<br>Medical Costs<br>Productivity Costs<br>Socioeconomic Deprivation<br>Socioeconomic Status |
| Quality of Life | Health Outcome<br>Health Status<br>Quality of Life |

By far the most commonly reported outcome among the included articles was quality of life (n=424), followed by return to work (n=179), social support (n=63), cost (n=50), and participation (n=41). Table 4 provides the five most common tools used to measure the 10 outcomes that are reported most. A table with the all the outcomes is provided in the Supplementary Materials (Table S5).

**Table 4.**
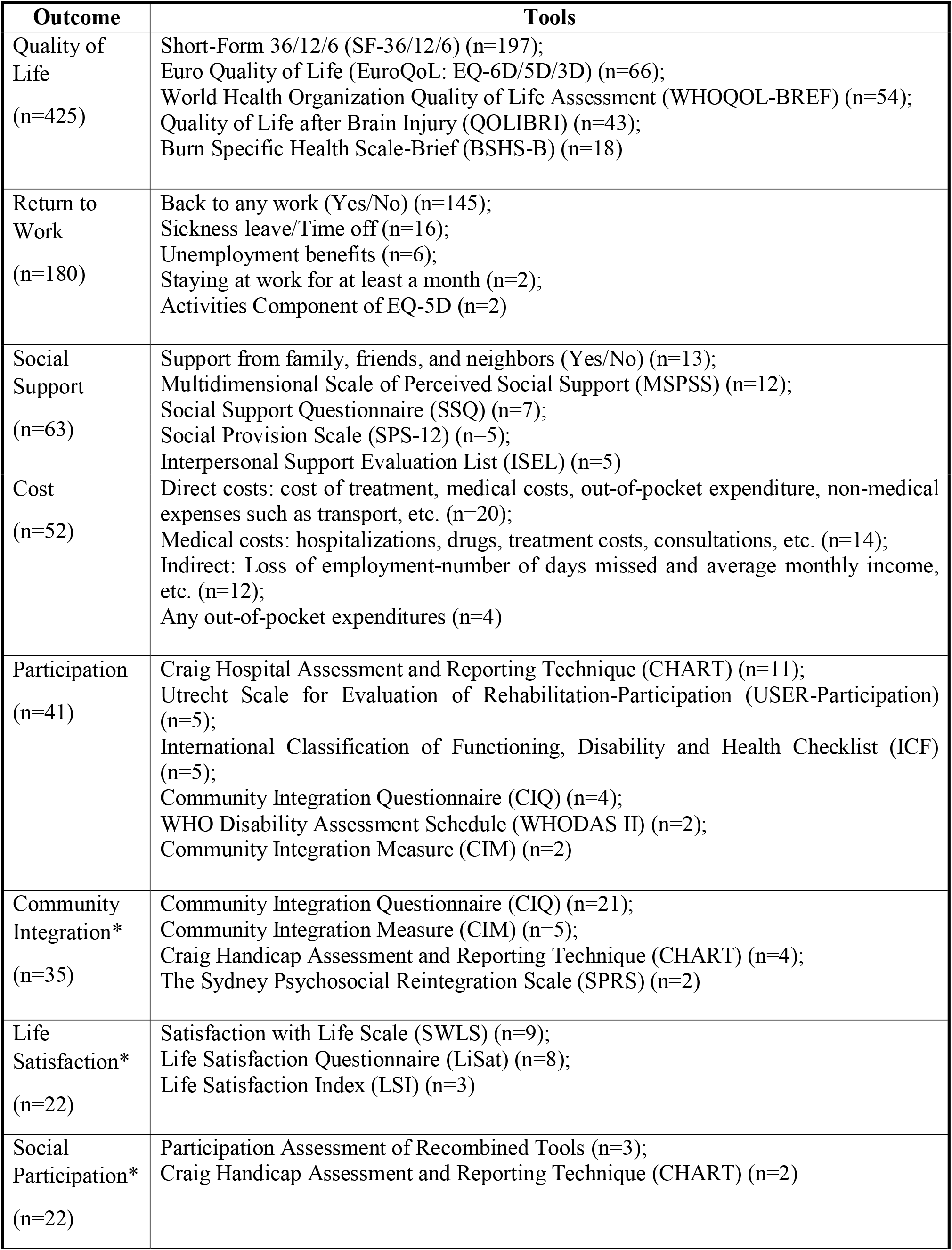
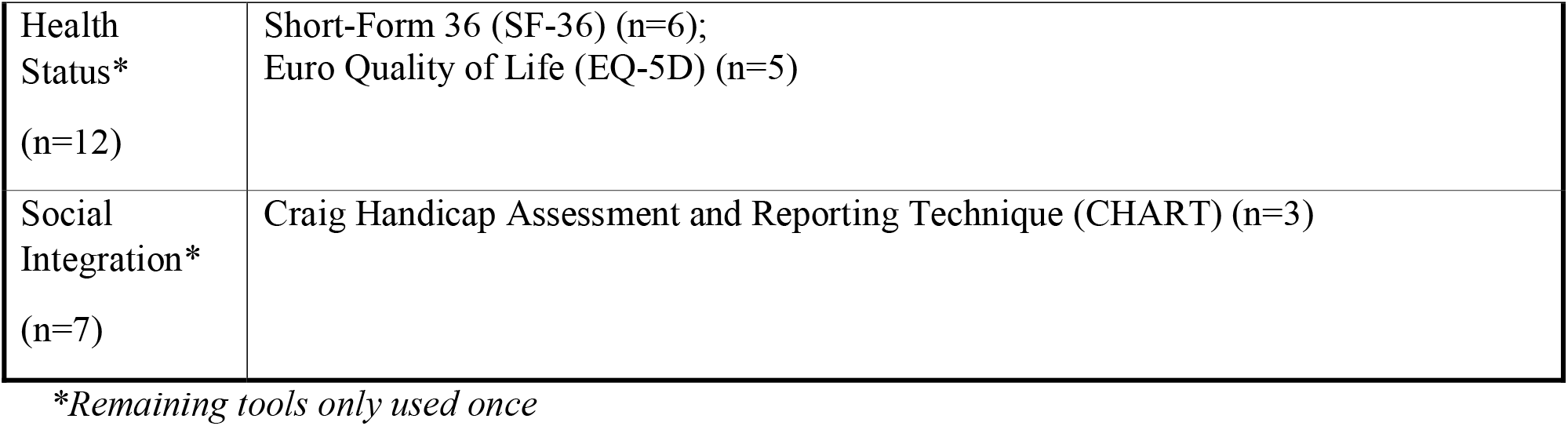
Most Reported Outcomes and Outcome Tools

On analyzing the five most reported outcomes among the three most common types of injury in both of the injury categories, it was seen that quality of life and return-to-work are the most commonly studied outcomes across all types of injuries (Table 5).

**Table 5.**
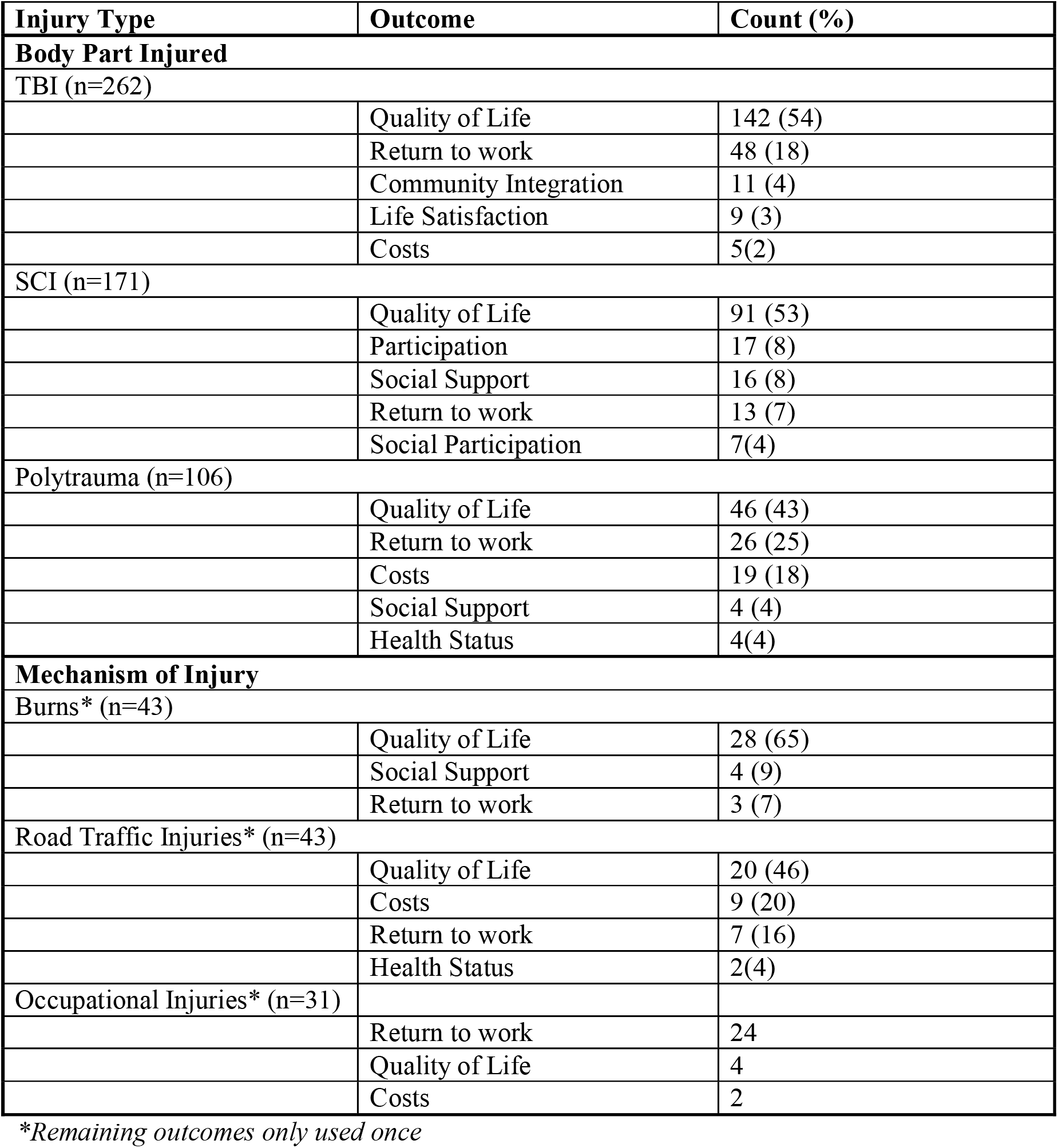
Most Reported Outcomes by Type of Injury

| **Injury Type** | **Outcome** | **Count (%)** |
| --- | --- | --- |
| **Body Part Injured** |  |  |
| TBI (n=262) |  |  |
|  | Quality of Life | 142 (54) |
|  | Return to work | 48 (18) |
|  | Community Integration | 11 (4) |
|  | Life Satisfaction | 9 (3) |
|  | Costs | 5(2) |
| SCI (n=171) |  |  |
|  | Quality of Life | 91 (53) |
|  | Participation | 17 (8) |
|  | Social Support | 16 (8) |
|  | Return to work | 13 (7) |
|  | Social Participation | 7(4) |
| Polytrauma (n=106) |  |  |
|  | Quality of Life | 46 (43) |
|  | Return to work | 26 (25) |
|  | Costs | 19 (18) |
|  | Social Support | 4 (4) |
|  | Health Status | 4(4) |
| **Mechanism of Injury** |  |  |
| Burns* (n=43) |  |  |
|  | Quality of Life | 28 (65) |
|  | Social Support | 4 (9) |
|  | Return to work | 3 (7) |
| Road Traffic Injuries* (n=43) |  |  |
|  | Quality of Life | 20 (46) |
|  | Costs | 9 (20) |
|  | Return to work | 7 (16) |
|  | Health Status | 2(4) |
| Occupational Injuries* (n=31) |  |  |
|  | Return to work | 24 |
|  | Quality of Life | 4 |
|  | Costs | 2 |

*\*Remaining outcomes only used once*

## 4. Discussion

### 4.1 Summary of Evidence

This scoping review was conducted to map the current knowledge on post-discharge socioeconomic and QoL outcomes in trauma patients by reviewing 78 outcomes extracted from 758 articles included in the study. The most commonly studied outcomes across the studies were quality of life, return-to-work (RTW), social support, cost, and participation. Short Form Survey (SF-36/12/8) and Craig Hospital Assessment and Reporting Technique (CHART) were the most commonly used tools to measure quality of life and participation, respectively. Simple yes or no answers were generally used to measure RTW and social support, while detailed questions on direct costs were used for estimating costs.

Our results show great variability in terms of the different socio-economic and QoL outcomes studied and measures or tools used. This highlights the high heterogeneity in the study of these outcomes across different types of trauma, patients, and settings. In the reviewed articles, a wide variety of different definitions and interpretations are used for the same outcomes. These inconsistences may be due to the lack of standardization of definitions of the outcomes. Despite the global guidelines such as ICF and WHODAS [27, 28] created to unify and standardize post-discharge outcomes, only a few studies have used them in defining or measuring outcomes after trauma.

This lacunae has also been pointed out in other reviews of post-discharge outcomes among trauma patients [29–31]. The variability makes comparisons across different types of injuries and settings challenging. There is increasing recognition of the need to standardize outcomes and outcome measures to improve estimation of risk, comparisons between settings, and delivery of care [32, 33]. Moving towards a global use of standardized outcome measures can build a body of homogeneous and uniform evidence on different outcomes. This may help future researchers and policy-makers to better understand the burden of post-discharge trauma.

Our results show that a substantial proportion (more than 80%) of the existing research on post-discharge patients comes from high-income settings. This is in line with research on orthopedic trauma [34], fractures [35], and TBI [36] that similarly show low trauma research outputs from LMIC settings. This means that the current knowledge on post-discharge outcomes in trauma patients is largely based on evidence from high-income settings [37–39].

In our analysis, in comparison to HICs, studies from LMICs were predominately cross-sectional in design without any cohort-based follow-ups (46% vs 71%). Such designs largely form the lower levels of evidence in health research hierarchy [40]. This indicates the need for more cohort-based research to study the trends and long-term effects of trauma morbidity, especially for LMIC settings. The reasons for this limited research from LMICs are attributed to the paucity of resources in these settings to conduct cohort follow-ups as well as the financial barriers in publishing them in peer-reviewed journals, which are often behind paywalls [38, 41].

On further analysis, our results show that more than half of the higher levels of evidence studies (prospective cohorts and RCTs) from LMICs were the product of collaborations with HIC institutions (9 out of 17). Therefore, future research projects could aim to foster such joint research to build local capacity, provide more resources, and increase the output of evidence from LMICs. Academic collaborations between HICs and LMICs has been suggested as the way forward to address this disparity in good quality research from LMICs which have disproportionate burden of trauma [42, 43].

Building on our findings, future research could narrowly focus of specific socioeconomic and QoL outcomes, and provide more detailed evidence on their definitions, measurements, and quality of evidence across different injuries and settings. Systematic reviews and meta-analyses of the outcomes based on this scoping review will enable in-depth study and comparisons of the outcomes across different studies. Additionally, other unexplored outcomes such as physiological functioning, psychological outcomes, and impact on caregivers can become the focus of future studies.

Future research on post-discharge socioeconomic outcomes in trauma patients should refer to international frameworks such as ICF and WHODAS to use the appropriate terminology and clearly define the outcome of interest. Using validated tools, which have been applied in different settings, to measure outcomes can build a standardized system of evidence to better understand the long-term outcomes of trauma. Additionally, future research can focus on outcomes such as social functioning, social roles, productivity costs, and catastrophic expenditure, which remain less studied in trauma patients.

### 4.2 Limitations

Our scoping review has several limitations. Firstly, the nature of scoping reviews is to focus on providing breadth rather than depth of information. But this was apt for the objective of this study to map the current knowledge on post-discharge outcomes in trauma patients. Secondly, our search was limited to studies in English, potentially leading to exclusion of relevant articles in other languages. Thirdly, the searches for the outcomes were based on terms from the ICF, the WHO Manual, and other articles accessed by the authors.

Given the vast variability in the outcomes reported, this could have excluded certain other socioeconomic and QoL outcomes. While the World Bank’s categorization of countries was from one point in time (i.e., 2018), the studies were from a 10-year period, during which time a country’s income categorization could have fluctuated. The articles mainly focused on patients with specific body type of injuries such as TBI or spinal cord injury. Therefore, it is difficult to comment on whether specific mechanisms of injury such as self-harm or falls, the second and third leading causes of injuries respectively, were adequately covered [4].

## 5. Conclusions

There is great variability in the outcomes and outcome measures reported across different types of injuries and settings. This would make comparative research and meta-analysis of the outcomes challenging. Quality of life, return-to-work, social support, cost, and participation were the main outcomes studied in post-discharge trauma patients. Post-discharge trauma studies should move towards building evidence based on standardized measurement of outcomes.

## Data Availability

Search strategy included in supplementary materials

## Declarations

This study was scoping review and did not involve participants directly, therefore did not require in ethics approval and consent.

### Authors’ contributions

All authors contributed to the study conception and design of the study. Data search, screening, extraction, and analysis were performed by Siddarth David and Martin Gerdin Wärnberg. The first draft of the manuscript was written by Siddarth David and all authors commented on previous versions of the manuscript. All authors read and approved the final manuscript.

## Acknowledgments

The authors would like to express their gratitude to all the members of the Thursday Truth Seekers, Mumbai for their support and feedback in shaping the paper. The authors would also like to express their gratitude to Anindra Z. Siqueira for proof-reading and editing the paper.

