## Supplemental Materials for "Measuring post-discharge socioeconomic and quality of life outcomes in trauma patients: A scoping review"

### Supplementary Materials

**Search Strategy**

A. Date Searched:

MEDLINE: 1^st^ August 2018

EMBASE: 1^st^ August 2018

Cochrane Library: 1^st^ August 2018

Web of Science: 2^nd^ August 2018

BASE: 2^nd^ August 2018

Global Index Medicus: 2^nd^ August 2018

B. Language:

English

C. Search Terms:
1. *MEDLINE*

((socio-economic[All Fields] OR social[All Fields] OR ("social adjustment"[MeSH Terms] OR ("social"[All Fields] AND "adjustment"[All Fields]) OR "social adjustment"[All Fields] OR ("social"[All Fields] AND "functioning"[All Fields]) OR "social functioning"[All Fields]) OR ("community participation"[MeSH Terms] OR ("community"[All Fields] AND "participation"[All Fields]) OR "community participation"[All Fields]) OR ("return to work"[MeSH Terms] OR ("return"[All Fields] AND "work"[All Fields]) OR "return to work"[All Fields]) OR ("economics"[MeSH Terms] OR "economics"[All Fields] OR "economic"[All Fields]) OR ("economics"[Subheading] OR "economics"[All Fields] OR "cost"[All Fields] OR "costs and cost analysis"[MeSH Terms] OR ("costs"[All Fields] AND "cost"[All Fields] AND "analysis"[All Fields]) OR "costs and cost analysis"[All Fields]) OR (out[All Fields] AND pocket[All Fields]) OR ("health expenditures"[MeSH Terms] OR ("health"[All Fields] AND "expenditures"[All Fields]) OR "health expenditures"[All Fields] OR "expenditure"[All Fields]) OR ("quality of life"[MeSH Terms] OR ("quality"[All Fields] AND "life"[All Fields]) OR "quality of life"[All Fields]) OR QOL[All Fields]) AND post[All Fields]) AND (("injuries"[Subheading] OR "injuries"[All Fields] OR "trauma"[All Fields] OR "wounds and injuries"[MeSH Terms] OR ("wounds"[All Fields] AND "injuries"[All Fields]) OR "wounds and injuries"[All Fields]) OR ("wounds and injuries"[MeSH Terms] OR ("wounds"[All Fields] AND "injuries"[All Fields]) OR "wounds and injuries"[All Fields] OR "traumatic injuries"[All Fields]) OR ("wounds and injuries"[MeSH Terms] OR ("wounds"[All Fields] AND "injuries"[All Fields]) OR "wounds and injuries"[All Fields] OR "injury"[All Fields]))

2. *EMBASE*

('injury'/exp AND 'hospital discharge' AND ('social aspects and related phenomena'/exp OR 'economic aspect'/exp OR 'health economics'/exp OR 'social status'/exp OR 'social behavior'/exp OR 'quality of life'/exp)) AND ([adolescent]/lim OR [adult]/lim OR [aged]/lim OR [middle aged]/lim OR [very elderly]/lim OR [young adult]/lim) AND 'human'/de

3. *Cochrane Review*

(("postdischarge"):ti,ab,kw or ("post-discharge"):ti,ab,kw or MeSH descriptor: [Patient Discharge]) and (("socioeconomic"):ti,ab,kw or ("socio-economic factors"):ti,ab,kw or ("socio-economic"):ti,ab,kw or ("socio-economic status"):ti,ab,kw or ("socio-economical"):ti,ab,kw or ("social function"):ti,ab,kw or ("social functioning"):ti,ab,kw or ("social functioning scale"):ti,ab,kw or ("social factor"):ti,ab,kw or ("economic factor"):ti,ab,kw or ("Quality of Life Enjoyment and Satisfaction Questionnaire"):ti,ab,kw or ("QOL"):ti,ab,kw or ("Quality of Life Index"):ti,ab,kw or MeSH descriptor: [Socioeconomic Factors] or MeSH descriptor: [Health Care Costs] or MeSH descriptor: [Quality of Life] or MeSH descriptor: [Costs and Cost Analysis]) and ((injury):ti,ab,kw or ("trauma and injury severity score"):ti,ab,kw or MeSH descriptor: [Wounds and Injuries]) and

4. *Web of Science*

TI= (("socio-economic" OR social OR "social functioning" OR "community participation" OR "return to work" OR economic OR cost* OR "out of pocket" OR expenditure* OR "quality of life" OR QOL) AND ("post discharge" AND trauma OR traumatic injur* OR injur*))Indexes: SCI-EXPANDED, SSCI, A&HCI, CPCI-S, CPCI-SSH, ESCI.

5. *BASE*

discharge and injury and quality-of-life or socioeconomic

6. *Global Medicus*

tw:((tw:(discharge)) AND (tw:(injury)) AND (tw:(socioeconomic)) OR (tw:(quality of life))) AND (mj:("Wounds and Injuries") AND limit:("humans" OR "adult")

**Table S1: Filled Prisma Scoping Review Checklist (ScR Checklist)**

| **Section/topic** | **#** | **Checklist item** | **Reported on page #** |
| --- | --- | --- | --- |
| **TITLE** | | | |
| Title | 1 | Identify the report as a scoping review. | 1 |
| **ABSTRACT** | | | |
| Structured summary | 2 | Provide a structured summary that includes (as applicable) background, objectives, eligibility criteria, sources of evidence, charting methods, results, and conclusions that relate to the review questions and objectives. | 1 |
| **INTRODUCTION** | | | |
| Rationale | 3 | Describe the rationale for the review in the context of what is already known. Explain why the review questions/objectives lend themselves to a scoping review approach. | 1-2 |
| Objectives | 4 | Provide an explicit statement of the questions and objectives being addressed with reference to their key elements (e.g., population or participants, concepts, and context) or other relevant key elements used to conceptualize the review questions and/or objectives. | 2 |
| **METHODS** | | | |
| Protocol and Registration | 5 | Indicate whether a review protocol exists; state if and where it can be accessed (e.g., a Web address); and, if available, provide registration information, including the registration number. | 2 |
| Eligibility Criteria | 6 | Specify characteristics of the sources of evidence used as eligibility criteria (e.g., years considered, language, and publication status), and provide a rationale. | 2 |
| Information Sources | 7 | Describe all information sources in the search (e.g., databases with dates of coverage and contact with authors to identify additional sources), as well as the date on which the most recent search was executed. | 2-3 |
| Search Strategy | 8 | Present the full electronic search strategy for at least 1 database, including any limits used, such that  it could be repeated. | 3 |
| Selection of Sources of Evidence | 9 | State the process for selecting sources of evidence (i.e., screening and eligibility) included in the scoping review. | 3 |
| Data Charting Process | 10 | Describe the methods of charting data from the included sources of evidence (e.g., calibrated forms or forms that have been tested by the team before their use, and whether data charting was done independently or in duplicate), and any processes for obtaining and confirming data from investigators. | 3 |
| Data Items | 11 | List and define all variables for which data were sought and any assumptions and simplifications made. | 3 |
| Critical Appraisal of Individual Sources of Evidence | 12 | If done, provide a rationale for conducting a critical appraisal of included sources of evidence; describe the methods used and how this information was used in any data synthesis (if appropriate). | 3 |
| Summary Measures | 13 | Not applicable for scoping reviews. | -- |
| Synthesis of Results | 14 | Describe the methods of handling and summarizing the data that were charted. | 3 |
| Risk of Bias Across Studies | 15 | Not applicable for scoping reviews. | -- |
| Additional Analyses | 16 | Not applicable for scoping reviews. | -- |
| **RESULTS** | | | |
| Selection of Sources of Evidence | 17 | Give numbers of sources of evidence screened, assessed for eligibility, and included in the review, with reasons for exclusions at each stage, ideally using a flow diagram. | 3-4 |
| Characteristics of Sources of Evidence | 18 | For each source of evidence, present characteristics for which data were charted and provide the citations. | 3-6 |
| Critical Appraisal Within Sources of Evidence | 19 | If done, present data on critical appraisal of included sources of evidence (see item 12). | NA |
| Results of Individual Sources of Evidence | 20 | For each included source of evidence, present the relevant data that were charted that relate to the review questions and objectives. | NA |
| Synthesis of Results | 21 | Summarize and/or present the charting results as they relate to the review questions and objectives. | 6-10 |
| Risk of Bias Across Studies | 22 | Not applicable for scoping reviews. | -- |
| Additional Analyses | 23 | Not applicable for scoping reviews. | -- |
| **DISCUSSION** | | | |
| Summary of Evidence | 24 | Summarize the main results (including an overview of concepts, themes, and types of evidence available), link to the review questions and objectives, and consider the relevance to key groups. | 10-12 |
| Limitations | 25 | Discuss the limitations of the scoping review process. | 12 |
| Conclusions | 26 | Provide a general interpretation of the results with respect to the review questions and objectives, as well as potential implications and/or next steps. | 12 |
| **FUNDING** | 27 | Describe sources of funding for the included sources of evidence, as well as sources of funding for the scoping review. Describe the role of the funders of the scoping review. | 13 |

*Tricco AC, Lillie E, Zarin W, Brien KKO, Colquhoun H, Levac D, et al.* *Research And Reporting Methods Prisma Extension for Scoping Reviews ( PRISMA-ScR ): Checklist and Explanation. Ann Intern Med. 2016;169(7):467–73*

**Table S2: Distribution of Included Studies by Year of Publication**

Distribution of included studies by year of publication

Number of Publications

|  | Number of Publications |
| --- | --- |
| 2009 | 38 |
| 2010 | 70 |
| 2011 | 63 |
| 2012 | 78 |
| 2013 | 60 |
| 2014 | 88 |
| 2015 | 88 |
| 2016 | 120 |
| 2017 | 111 |
| 2018 | 42 |

**The search period was from Aug 2009 to Aug 2018*

**Table S3: Distribution of Included Studies Which Had Follow-up Period**

Distribution of included studies which had a follow-up period

Frequency

|  | Frequency |
| --- | --- |
| Upto 1 month | 16 |
| 1-3 months | 29 |
| 4-6 months | 55 |
| 7-12 months | 180 |
| 13-24 months | 63 |
| 25-60 months | 98 |
| 61-120 months | 61 |
| More than 120 months | 72 |

**Table S4: Distribution of Included Studies by Country**

| **Country** | **Income** | **Frequency** |
| --- | --- | --- |
| USA | High | 162 |
| Australia | High | 97 |
| Netherlands | High | 52 |
| Canada | High | 41 |
| Multiple HICs | High | 37 |
| UK | High | 34 |
| Sweden | High | 28 |
| Taiwan | High | 26 |
| Different income countries | Mixed | 22 |
| Germany | High | 20 |
| Norway | High | 20 |
| Switzerland | High | 19 |
| France | High | 17 |
| China | Upper Middle | 14 |
| India | Lower Middle | 14 |
| Iran | Upper Middle | 14 |
| Brazil | Upper Middle | 11 |
| Italy | High | 9 |
| Japan | High | 9 |
| New Zealand | High | 9 |
| Malaysia | Upper Middle | 7 |
| South Korea | High | 7 |
| Spain | High | 7 |
| Denmark | High | 6 |
| Vietnam | Lower Middle | 6 |
| Bangladesh | Lower Middle | 5 |
| Finland | High | 5 |
| Pakistan | Lower Middle | 5 |
| Turkey | Upper Middle | 5 |
| Colombia | Upper Middle | 4 |
| Serbia | Upper Middle | 4 |
| Belgium | High | 3 |
| Indonesia | Lower Middle | 3 |
| Israel | High | 3 |
| Mexico | Upper Middle | 3 |
| Poland | High | 3 |
| Thailand | Upper Middle | 3 |
| Estonia | High | 2 |
| Greece | High | 2 |
| Ireland | High | 2 |
| Multiple LMICs | Lower Middle | 2 |
| Nepal | Low | 2 |
| Singapore | High | 2 |
| Austria | High | 1 |
| Benin | Low | 1 |
| Chile | High | 1 |
| Egypt | Lower Middle | 1 |
| Haiti | Low | 1 |
| Lithuania | High | 1 |
| Malawi | Low | 1 |
| Morocco | Upper Middle | 1 |
| Nigeria | Lower Middle | 1 |
| Romania | Upper Middle | 1 |
| Sri Lanka | Lower Middle | 1 |
| Uganda | Low | 1 |

**Table S5: List of Reported Outcomes and Outcome Measures**

| **Outcome** | **Measure** |
| --- | --- |
| Quality of Life  (n=424) | Short-Form 36/12/6 (SF-36/12/6) (n=197)  Euro Quality of Life (EuroQoL: EQ-6D/5D/3D) (n=66)  World Health Organization Quality of Life Assessment (WHOQOL-BREF) (n=54)  Quality of Life after Brain Injury (QOLIBRI) (n=43)  Burn Specific Health Scale-Brief (BSHS-B) (n=18)  Sickness Impact Profile (SIP) (n=11)  Satisfaction with Life Scale (n=4)  Health Utility Index (n=3)  Assessment of Quality of Life (AQOL) (n=2)  Quality of Life Scale (QoLS) (n=2)  Clinical Tracking Form (n=2)  Quality of Well Being Self-Administered (QWB-SA) Scale (n=2)  Knee Injury and Osteoarthritis Outcome Score (KOOS)-QoL (n=2)  Reintegration to Normal Living Index (RNL)  Life Satisfaction Questionnaire-9 (LISAT-9)  Ferrans and Powers Quality of Life Index – Spinal Cord Injury Version  Open-ended questions: 1) How do you define QOL? and 2) Can you mention 5 things that are important for your QOL?  Disablement in the Physically Active Scale  Personal Well-Being Index (PWI)  QoL-Feedback from Hanssen-Doose and Schule  Nottingham Health Profile  Patient Reported Impact of Spasticity Measure (PRISM)  Quality of Life Profile for Adults with Physical Disabilities (QOLP-PD)  Sense of Well-Being Inventory (SWBI)  EUROHIS Quality of Life Scale  Patient Generated Index (PGI)  Life Satisfaction Index (LSI)  Quality of Life Index  Life Satisfaction Survey  Quality of Life Attribute (QLA)  Questions on Life Satisfaction  Dutch SCI Patient Organization Questionnaire  National Eye Institute 25-Item Visual Function Questionnaire (NEI VFQ-25)  International Osteoporosis Foundation Quality of Life Questionnaire (IOF QLQ), SF-36  Condition was stable, improved or decreased compared to the initial condition  Symptom Checklist-90-R (SCL-90-R)  PROMIS/Neuro-QOL, Spinal Cord Injury Quality of Life Measurement System (SCI-QOL) and TBI-QOL  Problem Checklist (PCL)  Life Situation Questionnaire-Revised  Functional Status Questionnaire (FSQ)  Disabilities of the Arm, Shoulder and Hand (DASH) score and the Ulm questionnaire  Perceived Quality of Life Scale (PQOL)  Having social support, caregiver, financial resources  PAECC (Project for the Epidemiological Analysis of Critical Care Patients) QOL (Quality of Life) questionnaire |
| Return to Work  (n=180) | Back to any work (Yes/No) (n=145)  Sickness leave/Time off (n=16)  Unemployment benefits (n=6)  Staying at work for at least a month (n=2)  Activities Component of EQ-5D (n=2)  4 categories: (0) previous work or study resumed; (1) previous work or study resumed, but with lower demands or part-time; (2) previous work or study not resumed, different work resumed on a significantly lower level; and (3) not working.  4 questions: (1) patients who could not return to their pre-injury jobs and work at pre-injury levels; (2) those who returned to pre-injury duties but were transferred to less-demanding positions; (3) those who returned to pre-injury duties but worked fewer hours and at reduced levels and (4) those who returned to pre-injury duties.  full-time or part-time gainful military or civilian employment  Worked at least 50% in at least a year after the injury  Had returned to pre-injury work or to a new job, with at least 20 hours per week at work, during 1 year  Return to part-time employment only (30 h/week).  Individuals working full-time (more than 37.5 h/week) or part-time (less than 27.5 h/week)  Duration of any interruption of work  ≥ 25% employment  Patient's estimate of return to work  Part-time, full-time or with modified work or light duties  Return-to-Work Self-Efficacy (RTWSE) scale  Self-efficacy: likelihood of your returning to work within one month?  ‘Did you have a paid job of any kind at the time of the accident?’, ‘Did you take any time off work as a result of your accident?’, ‘How long have you been back at work in this job?’, ‘Thinking about those jobs, did you work at any of them for three months or more?’, and ‘Of the jobs you had for three months or more, did you leave either/any of them for a reason related to your accident?’ |
| Social Support  (n=63) | Support from family, friends, and neighbors (Yes/No) (n=13)  Multidimensional Scale of Perceived Social Support (MSPSS) (n=12)  Social Support Questionnaire (SSQ) (n=7)  Social Provision Scale (SPS-12) (n=5)  Interpersonal Support Evaluation List (ISEL) (n=3)  Social Support Scale (n=2)  Perceived Social Support (PSS) (n=2)  Social Support Index  Personal Resource Questionnaire  National Institute of Health (NIH) Social Relationship Scales  Medical Outcomes Study Social Support Survey (MOS-SSS)  ICF  Inclusive, supportive, accepting of the situation, abandonment  DRRI-Post deployment Social Support Scale (PDSS)  The Sydney Psycho-social Reintegration Survey (SPRS)  Abbreviated Duke Social Support Index (ADSSI)  Involvement of family and friends  Discharged Burn Patients’ Social Support inventor  Family awareness, understanding and support  Schuster Social Support Questions (SSSQ)  Provided Social Support Scale (PSSS)  Behavioral Risk Factor Surveillance System  Goldsmith Social Support Scale  Social support, social relationship  Crisis Support Scale (CSS)  Source of Social Support Inventory (SSSI)  Provision of Social Relationship (PSR) |
| Cost  (n=52) | Direct costs: cost of treatment, medical costs, out-of-pocket expenditure, non-medical expenses such as transport, etc. (n=20)  Medical costs: hospitalizations, drugs, treatment costs, consultations, etc. (n=14)  Indirect: Loss of employment-number of days missed and average monthly income, etc. (n=12)  Any out-of-pocket expenditure (n=4)  Mean Hospital charges  Catastrophic out-of-pocket expenditure is 30% of spending  Catastrophic out-of-pocket expenditure is 40% of spending  Borrow, sell assets  Monthly income before and after the injury  Spending on medical care to the loss of income, loss of resources, borrowing  Mean labor cost was applied to the length of stay  Loss of productivity: days of job missed and daily salary  Loss of employment, work absences, leave by family members |
| Participation  (n=41) | Craig Hospital Assessment and Reporting Technique (CHART) (n=11)  Utrecht Scale for Evaluation of Rehabilitation-Participation (USER-Participation) (n=5)  International Classification of Functioning, Disability and Health checklist (ICF) (n=5)  Community Integration Questionnaire (CIQ) (n=4)  WHO Disability Assessment Schedule (WHODAS II) (n=2)  Community Integration Measure (CIM) (n=2)  Sickness Impact Profile  Rivermead Head Injury Follow-up Questionnaire  Adult Subjective Assessment of Participation (ASAP):  Return to their usual social activities  Nottwil Environmental Factors Inventory  International Spinal Cord Injury (SCI) Activities and Participation (A&P) Basic Data Set  Employment Status  Inability to perform life roles and attend activities/hobbies  Earning income and work, being useful to others, community participation, and having skills and knowledge |
| Community Integration  (n=35) | Community Integration Questionnaire (CIQ) (n=21)  Community Integration Measure (CIM) (n=5)  Craig Handicap Assessment and Reporting Technique (CHART) (n=4)  The Sydney Psychosocial Reintegration Scale (SPRS) (n=2)  PARTS/M  ICF  Cannot leave the home  Participation in work, school, home, and other productive and meaningful activities; participation in social relationships and activities; and the ability to live independently (Community Reintegration for Service Member instrument-CRIS) |
| Social Participation  (n=22) | Participation Assessment of Recombined Tools (n=3)  Craig Handicap Assessment and Reporting Technique (CHART) (n=2)  Life Impact Burn Recovery Evaluation (LIBRE) Profile  Social Support Questionnaire  Community Integration Questionnaire (CIQ-SI)  Support of family, friends. Motivation and encouragement  IMPACT-S  Measure of social fit  Life Satisfaction Questionnaire (LiSat-9)  Comfort interacting with family and friends  Sydney Psychosocial Reintegration Scale (SPRS)  PART-O Social Relations sub-scale  Adult Subjective Assessment of Participation (ASAP)  Past year, participation in the following independent activities: hobby, health/sport, production/work, education/culture, social climate improvement, safety management, welfare, community event, and no participation  (I) Gone to the movies, concerts, plays, or sporting events; (ii) gone to fairs, museums, or exhibits; (iii) attended meetings, appointments, classes, or lectures; (iv) gone to church or temple services; (v) gone on pleasure drives or picnics; (vi) played cards, bingo etc. with others; (vii) gone to family/friends’ homes for a meal; (viii) participated in active sports or swimming; (ix) worked in the garden/yard or at a hobby and (x) performed community or volunteer work. |
| Life Satisfaction  (n=22) | Satisfaction with Life Scale (SWLS) (n=9)  Life Satisfaction Questionnaire (LiSat) (n=8)  LSI-Life Satisfaction Index (n=3) |
| Health Status  (n=12) | Short-Form 36 (SF-36) (n=6);  Euro Quality of Life (EQ-5D) (n=5)  ‘Overall, how would you rate your health during the past month?’’ A: ‘‘poor’’, ‘‘fair’’, ‘‘good’’, ‘‘very good’’, or ‘‘excellent.’ |
| Social Integration  (n=7) | Craig Handicap Assessment and Reporting Technique (CHART) (n=3)  Community Integration Questionnaire (CIQ)  Rivermead Head Injury Follow-Up Questionnaire (RHFUQ)  World Health Organization Disability Assessment Schedule II (WHODAS II)  Access to social activities, residential programs, and leisure activities in a specialized or community environment |
| Independence  (n=6) | Barthel Index of Activity in Daily Life (n=3)  Houghton Scale score  ICF  Functional Independence Measure (FIM) |
| Social Functioning  (n=6) | SF-36 (n=3)  Glasgow Outcome Scale  Sydney psychosocial reintegration scale (SPRS)  Social isolation and decreased participation in activities |
| Financial Coping/Source  (n=5) | Using savings, borrowing from others or selling assets (n=2)  Borrow money from relatives/friends, reduced consumption of basic goods, reduced food consumption, took loans, sold assets, other  Borrowing, selling assets, catastrophic spending (more than 20% of household income and 40% household income)  Lost assets, reduced spending, borrowing |
| Mobility  (n=5) | Timed up-and-go test (length of time taken to rise from a chair, walk 3 meters, return to the chair and sit back down: poor mobility = 20+ seconds); gait test (time and number of steps in the faster of 2 trials of walking 4 meters); also mobility categories ranging from totally immobile to able to walk without aids) (n=2)  Groningen Activity Restriction Scale  Able to walk for 6 minutes  Walk Test |
| Employment Outcomes  (n=4) | Returned to work (Yes/No) (n=3)  Remained employed for 1 year after placement, gaining employment that lasted at least 90 days |
| Employment Status  (n=4) | Yes, No (n=3)  Paid employment for 1 hour a week or mor |
| Social Life  (n=4) | Limiting social network/contacts, giving up hobbies/activities of leisure  Quality of Life Interview-Brief Version (QOLI-BV)  Yes, No  Glasgow Outcome Scale Extended |
| Medical Costs  (n=3) | Consultation, medical procedures, OPD costs, nursing costs, bed charges, ICU costs, physiotherapy, transportation  Emergency, inpatient, outpatient, pharmacy, radiology, post-acute care, and other (post-discharge services)  Out-of-pocket in OPD and IPD |
| Satisfaction/Satisfaction with life  (n=3) | Satisfaction with Life Scale (n=3) |
| Sentinel Events  (n=3) | Sentinel Events Questionnaire (n=3) |
| Social Interactions  (n=3) | Participation in peer activities and events  Evaluation of Social Interaction (ESI)  Not participate in community activities: church, weddings, events; limited interaction with family and friends |
| Social Isolation  (n=3) | Social Isolation Index  UCLA Loneliness Scale (Version 3)  Life Situation Questionnaire (LSQ) |
| Social Roles  (n=3) | Fulfilling gender roles  Duties as parents, authority as parents  Taking up opposite gender roles |
| Support  (n=3) | Professional help, advocacy, information, community resources, family, and friends  ICF  Organizational Support |
| Community and Social Life  (n=2) | ICF (n=2) |
| Daily Activity  (n=2) | Barthel Index  Burn Specific Health Scale-Brief (BSHS-B) |
| Environmental barriers  (n=2) | Craig Hospital Inventory of Environmental Factors-Short Form (CHIEF-SF)  (n=2) |
| Financial Difficulty/Load  (n=2) | Yes, No  0–100 scale: 100 refers to losing all of the family’s savings (loans, selling properties); 75 means spending all of the family’s savings; 50 refers to using all the monthly income of the family; 25 means more than half of the monthly income of the family is consumed; 0 means there was no charge |
| Physical Activity  (n=2) | Physical Activity Recall Assessment for People with Spinal Cord Injury (PARA-SCI)  The Physical Activity Scale for Individuals with Physical Disabilities (PASIPD): lawn and garden work, housework, vigorous sport and recreation, moderate sport and recreation, and occupation and transportation |
| Reintegration  (n=2) | Reintegration to Normal Living (RNL) Index  Sydney Psychosocial Reintegration Scale (SPRS) |
| Social Adjustment  (n=2) | Social involvement and perceived social support  Social Adjustment Scale (SAS) |
| Social Inclusion  (n=2) | Integration; Participation; Support  Institut Guttmann social scale |
| Time off work  (n=2) | Months (n=2) |
| Work  (n=2) | Burn Specific Health Scale-Brief (BSHS-B)  Full-time work (more than 30 h/week) or part-time work (less than 30 h/week) |
| Work-place support  (n=2) | Supervisor, co-worker, work environment  Yes, No |
| Social Activities  (n=1) | Social/Role Activities Limitations |
| Productivity Costs  (n=1) | 30% of wages deducted for illness absenteeism |
| Activities  (n=1) | Household chores, running the house |
| Caregivers  (n=1) | Family, friends |
| Economic Impact  (n=1) | Personal income, standard of living and household income adequacy (household income was adequate to meet every day needs such as accommodation, food and clothing was asked using a four-point scale: More than enough, enough, just enough and not enough (grouped as ‘more than enough/enough’; ‘just enough/not enough’) |
| Environmental Assistance  (n=1) | Assistance at home |
| Everyday Activities  (n=1) | Independent living |
| Family Life  (n=1) | Changes in the roles and responsibilities of different members within the family |
| Family Support  (n=1) | Emotional, informational, tangible, affectionate support and positive social interaction from family members |
| Financial Security  (n=1) | Not meeting basic family needs, relying on others for assistance, and feeling helpless because they could not work or contribute in their usual role |
| Financial status  (n=1) | Having insufficient money to support oneself and/or family, and provide for one’s health and rehabilitation needs |
| Financial Strain  (n=1) | Having to borrow money or goods greater than $100 in value or accessing funds via superannuation accounts, charitable organizations, savings plans, or via sale of assets |
| Health Outcome  (n=1) | EQ-5D |
| Impact on employment or studies  (n=1) | Yes, No |
| Impact on familial or affective life  (n=1) | Yes, No |
| Impact on leisure or sport activities  (n=1) | Yes, No |
| Independent Functioning  (n=1) | Barthel Index |
| Integration  (n=1) | Sydney Psychosocial Reintegration Scale (SPRS) |
| Interpersonal Relationships  (n=1) | Burn Specific Health Scale-Brief (BSHS-B) |
| Job stability  (n=1) | More than 1 employer |
| Labour Market Participation  (n=1) | Paid employment |
| Life Participation  (n=1) | Participation Assessment with Recombined Tools Objective (PART-O). All were scored based on the number of times per month a participant had engaged in the activity. For each item, those below the 25th percentile were scored as 0, those between the 25th and 75th percentiles were scored as 1, and those above the 75th percentile were scored as 2. The scores for all four items were then summed (range 0 – 8) and categorized into Low participation (summed score 0 –1), Medium participation (summed score 2– 4), and High participation (summed score 5– 8). |
| Limited Social Interaction  (n=1) | Inability to participate in social events |
| Participate in social life  (n=1) | Modified Branholms Interest Checklist |
| Perceived Social Support  (n=1) | 1) I have enough friends and social life, (2) I have close contact with members of my family, and (3) When things get really bad, I know I can count on my friends and family for help |
| Restricted Mobility  (n=1) | Inability to eat, move around independently |
| Return to leisure activities  (n=1) | Return to leisure activities, community events |
| Return to Work costs  (n=1) | Time spent in the hospital, work day lost and their average monthly salary of patients and attendants |
| Role of Caregiver  (n=1) | Craig Handicap Assessment and Reporting Technique |
| Social Adaptation  (n=1) | Social Adaptation Self Evaluation Scale (SASS) |
| Social Behavior  (n=1) | Sickness Impact Profile |
| Social Consequences  (n=1) | Effect on family, work and social life |
| Social Costs  (n=1) | Individual costs (income, medical costs), family costs (loss of resources, police/legal costs), community costs (loss of businesses, infrastructure, housing prices) |
| Social Leisure Activities  (n=1) | Shortened Version of the Nottingham Leisure Questionnaire (NLQ) |
| Social Networks  (n=1) | Social Network Index (SNI) |
| Social Role Change  (n=1) | Changes in family role |
| Social Role Participation  (n=1) | Work, volunteering, recreation and leisure pursuits, and social and family role function |
| Socioeconomic deprivation  (n=1) | Welsh Index of Multiple Deprivation (income, employment, health, education, access to services, housing, physical environment and community safety) |
| Socioeconomic status  (n=1) | Ecological indices of social and material deprivation (Raymond & Pamplon 2012) |
| Sustained Return to Work  (n=1) | Back at work for >28 days |
| Unpaid work  (n=1) | Shopping, household chores, caring for children (Health and Labor Questionnaire) |
| Work Satisfaction  (n=1) | Yes, No |
